# COVID-19 Antibody Seroprevalence in Santa Clara County, California

**DOI:** 10.1101/2020.04.14.20062463

**Authors:** Eran Bendavid, Bianca Mulaney, Neeraj Sood, Soleil Shah, Emilia Ling, Rebecca Bromley-Dulfano, Cara Lai, Zoe Weissberg, Rodrigo Saavedra-Walker, Jim Tedrow, Dona Tversky, Andrew Bogan, Thomas Kupiec, Daniel Eichner, Ribhav Gupta, John P.A. Ioannidis, Jay Bhattacharya

## Abstract

**Background:** Addressing COVID-19 is a pressing health and social concern. To date, many epidemic projections and policies addressing COVID-19 have been designed without seroprevalence data to inform epidemic parameters. We measured the seroprevalence of antibodies to SARS-CoV-2 in a community sample drawn from Santa Clara County.

**Methods:** On April 3-4, 2020, we tested county residents for antibodies to SARS-CoV-2 using a lateral flow immunoassay. Participants were recruited using Facebook ads targeting a sample of individuals living within the county by demographic and geographic characteristics. We estimate weights to adjust our sample to match the zip code, sex, and race/ethnicity distribution within the county. We report both the weighted and unweighted prevalence of antibodies to SARS-CoV-2. We also adjust for test performance characteristics by combining data from 16 independent samples obtained from manufacturer’s data, regulatory submissions, and independent evaluations: 13 samples for specificity (3,324 specimens) and 3 samples for sensitivity (157 specimens).

**Results:** The raw prevalence of antibodies to SARS-CoV-2 in our sample was 1.5% (exact binomial 95CI 1.1-2.0%). Test performance specificity in our data was 99.5% (95CI 99.2-99.7%) and sensitivity was 82.8% (95CI 76.0-88.4%). The unweighted prevalence adjusted for test performance characteristics was 1.2% (95CI 0.7-1.8%). After weighting for population demographics of Santa Clara County, the prevalence was 2.8% (95CI 1.3-4.7%), using bootstrap to estimate confidence bounds. These prevalence point estimates imply that 54,000 (95CI 25,000 to 91,000 using weighted prevalence; 23,000 with 95CI 14,000-35,000 using unweighted prevalence) people were infected in Santa Clara County by early April, many more than the approximately 1,000 confirmed cases at the time of the survey.

**Conclusions:** The estimated population prevalence of SARS-CoV-2 antibodies in Santa Clara County implies that the infection may be much more widespread than indicated by the number of confirmed cases. More studies are needed to improve precision of prevalence estimates. Locally-derived population prevalence estimates should be used to calibrate epidemic and mortality projections.

## Introduction

The first two cases of COVID-19 in Santa Clara County, California were identified in returning travelers on January 31 and on February 1, 2020, and the first COVID-19 death in the county was announced on March 9.^1^ In the following month, nearly 1,000 additional cases were identified in Santa Clara County, showing a pattern of rapid case increase reflective of community transmission as well as the scaling up of SARS-CoV-2 viral testing that was common across many communities globally. In some countries, the rapid increase in COVID-19 case counts and hospitalizations has overwhelmed health systems and led to large reductions in social and economic activities. The measures adopted to slow the spread of COVID-19 were justified by projected estimates of health care system capacity and case fatality rate. These projections suggested that, in the absence of strict measures to reduce transmission, the COVID-19 pandemic could overwhelm existing hospital bed and ICU capacity throughout the United States and lead to over 2 million deaths.^2^

Measuring fatality rates and projecting the number of deaths depend on estimates of the total number of infections. To date, in the absence of seroprevalence surveys, estimates of the fatality rate have relied on the number of confirmed cases multiplied by an estimated factor representing unknown or asymptomatic cases to arrive at the number of infections.^3–6^ However, the magnitude of that factor is highly uncertain. Because the implications of infection fatality rate and projected deaths are large, the extent of COVID-19 infection under-ascertainment (the multiplier used to arrive from cases to infections) has been a topic of great interest and provided estimates of the number of infections about 1-6-fold higher than the number of cases.^7–10^ The extent of infection under-ascertainment has been difficult to assess because of three biasing processes: (i) cases have been diagnosed with PCR-based tests, which do not provide information about resolved infections; (ii) the majority of cases tested early in the course of the epidemic have been acutely ill and highly symptomatic, while most asymptomatic or mildly symptomatic individuals have not been tested; and (iii) PCR-based testing rates have been highly variable across contexts and over time, leading to inaccurate relationships between the number of cases and infections. If, in the absence of interventions, the epidemic’s early doubling time is estimated to be four days, then by February 27th, 2020, when the third case was identified in Santa Clara County, the county may have already had 256 infections.^6,11,12^ And an autopsy report released on April 20 showed that the first death in the county occurred as early as February 6, suggesting that the virus may have been circulating in the community much earlier than recognized.^13^

At the time of this study, Santa Clara County had the largest number of confirmed cases of any county in Northern California. The county also had several of the earliest known cases of COVID-19 in the state - including one of the first presumed cases of community-acquired disease - making it an especially appropriate location to test a population-level sample for the presence of active and *past* infections.

On April 3rd and 4th, 2020 we conducted a survey of residents of Santa Clara County to measure the seroprevalence of antibodies to SARS-CoV-2 and better approximate the number of infections. Our goal is to provide new and well-measured data for informing epidemic models, projections, and public policy decisions.

## Methods

We conducted serologic testing for SARS-CoV-2 antibodies in 3,330 adults and children in Santa Clara County using capillary blood draws and a lateral flow immunoassay. In this section we describe our sampling and recruitment approaches, specimen collection methods, antibody testing procedure, test kit validation, and statistical methods. Our protocol was informed by a World Health Organization protocol for population-level COVID-19 antibody testing.^14^ We conducted our study with the cooperation of the Santa Clara County Department of Public Health. The Institutional Review Board at Stanford University approved the study prior to recruitment.

### Study Participants and Sample Recruitment

We recruited participants by placing targeted advertisements on Facebook aimed at residents of Santa Clara County. We used Facebook to quickly reach a large number of county residents and because it allows for granular targeting by zip code and sociodemographic characteristics.^15^ We posted our advertisements targeting two populations: ads aimed at a representative population of the county by zip code, and specially targeted ads to balance our sample for under-represented zip codes. In addition, we capped registrations from overrepresented areas after our registration slots filled up quickly with participants from wealthier zip codes. Individuals who clicked on the advertisement were directed to a survey hosted by the Stanford REDcap platform, which provided information about the study.^16^ The survey asked for six data elements: zip code of residence, age, sex, race/ethnicity, underlying co-morbidities, and prior clinical symptoms. Over 24 hours, we registered 3,285 adults, and each adult was allowed to bring one child from the same household with them (889 children registered). Additional details of the participant selection process are provided below (Additional Data and Response to Comments section below).

### Specimen Collection and Testing Methods

We established drive-through test sites in three locations spaced across Santa Clara County: two county parks in Los Gatos and San Jose, and a church in Mountain View. Only individuals with a participant ID were allowed into the testing area. Verbal informed consent was obtained to minimize participant and staff exposure. With participants in their vehicles, sample collectors in personal protective equipment drew 50-200µL of capillary blood into an EDTA-coated microtainer. Tubes were barcoded and linked with the participant ID. Samples were couriered from the collection sites to a test reading facility with steady lighting and climate conditions. Technicians drew whole blood up to a fill line on the manufacturer’s pipette and placed it in the test kit well, followed by a buffer. Test kits were read 12-20 minutes after the buffer was placed. Technicians barcoded tests to match sample barcodes and documented all test results.

### Test Kit Performance

The manufacturer’s performance characteristics were available prior to the study (using 85 confirmed positive and 371 confirmed negative samples). We conducted additional testing to assess the kit performance, and continued collecting information from assessments of test performance to incorporate into the analysis. Broadly, test performance was assessed against gold-standard positive specimens from patients with PCR-confirmed COVID-19 (with or without additional confirmation of antibody presence) for sensitivity, and gold-standard negative specimens from pre-COVID-era and early-COVID-era specimens. More detail on the data provenance, procedures to assess test performance characteristics, and concordance between gold standard and kit results is provided below (Additional Data and Response to Comments section).

### Statistical Analysis

Our estimation of the prevalence of COVID-19 proceeded in three steps. First, we report the raw frequencies of positive tests as a proportion of the final sample size. Second, we report the estimated sample prevalence, adjusted for test performance characteristics. Because SARS-CoV-2 lateral flow assays are new, we gathered all available information on test performance characteristics (sensitivity and specificity), with a focus on test specificity, which can be of paramount importance when prevalence is not high. We use an estimate of test sensitivity and specificity based on pooling all information available to us. Details of each sample, including test kit agreement numbers, specimen type, and available information on data provenance, is provided in the Additional Data and Response to Comments section.

Third, we report the weighted prevalence after weighting for the zip code, sex, and race/ethnicity (non-Hispanic White, Asian, Hispanic, and other) distributions of Santa Clara County (as measured in the 2018 American Community Survey). We chose these three adjustors because they contributed to the largest imbalance in our sample, and because including additional adjustors would result in small-N bins. Our weights were the zip-sex-race proportion in Santa Clara County divided by the zip-sex-race proportion in our sample, for each zip-sex-race combination in the county and in the sample. We show the distribution of our sample weights and provide more details in the Additional Data and Response to Comments section and in the Statistical Appendix.

We use a bootstrap procedure to estimate confidence bounds for the unweighted and weighted prevalence, while accounting for sampling error and propagating the uncertainty in the sensitivity and specificity. We use the basic percentile of the bootstrap distribution to construct confidence intervals.^17^ This procedure assumes that the community sample, negative control sample, and positive control sample are drawn independently. More details on our bootstrap procedures are in the Additional Data and Response to Comments section and in the Statistical Appendix.

## Results

The test kit used in this study (Premier Biotech, Minneapolis, MN) was tested prior to field deployment, and additional test performance data have been incorporated. In addition to the information we had prior to deployment (2 samples for specificity and 2 for sensitivity), we collected information on 12 additional samples used for assessing specificity of this particular kit (11 for specificity and 1 for sensitivity), for a total of 3,324 specimens tested for specificity and 157 specimens tested for sensitivity across all 16 samples. Additional details on the size and performance of each sample is provided in Additional Data and Response to Comments section. The combined specificity using data from all the samples was 99.5% (exact binomial 95CI 99.2-99.7%) and sensitivity was 82.8 (exact binomial 95CI 76.0-88.4%).

Our study included 3,439 individuals that registered for the study and arrived at testing sites. We excluded observations of individuals who could not be tested (e.g. unable to obtain blood or blood clotted, N=49), whose test results could not be matched to their personal data (e.g. if an incorrect participant ID was recorded onsite, N=30), who did not reside in Santa Clara County (N=29), and who had invalid test results (no Control band, N=1). This yielded an analytic sample of 3,330 individuals with complete records including survey registration, attendance at a test site for specimen collection, and lab results (Figure 1). The sample distribution meaningfully deviated from that of the Santa Clara County population along several dimensions: sex (63% in sample was female, 50% in county); race (8% of the sample was Hispanic, 26% in the county; 19% of the sample was Asian, 28% in the county); and zip distribution (median participant density, 1.6 per 1,000 population, IQR 0.9-3.6). Table 1 includes demographic characteristics of our unadjusted sample, population-adjusted sample, and Santa Clara County.^18^ Figure 2 shows the geographical zip code distribution of study participants in the county (counts and density per 1,000 population).

**Table 1:** Sample characteristics, relative to Santa Clara County population estimates from the 2018 American Community Survey

| <u>Characteristic</u> |  | <u>Sample –</u><br><u>unweighted</u> | <u>Sample –</u><br><u>weighted</u> | <u>County</u> |
| --- | --- | --- | --- | --- |
| Population (N) |  | 3,330 | 3,330 | 1,943,411 |
| Women (%) |  | 63.1 | 49.7 | 49.5 |
| Men (%) |  | 36.9 | 50.3 | 50.5 |
| Age (%) | 0-4 | 2.1 | 2.6 | 6.2 |
|  | 5-18 | 16.5 | 14.5 | 18.6 |
|  | 19-64 | 76.3 | 78.4 | 62.3 |
|  | ≥65 | 5.0 | 4.5 | 12.9 |
| Race/ethnicity (%) | Non-Hispanic white | 64.1 | 35.4 | 33.1 |
|  | Hispanic | 8.0 | 24.9 | 26.3 |
|  | Asian | 18.7 | 28.9 | 27.8 |
|  | Other | 9.2 | 10.8 | 12.8 |

**Figure 1:**
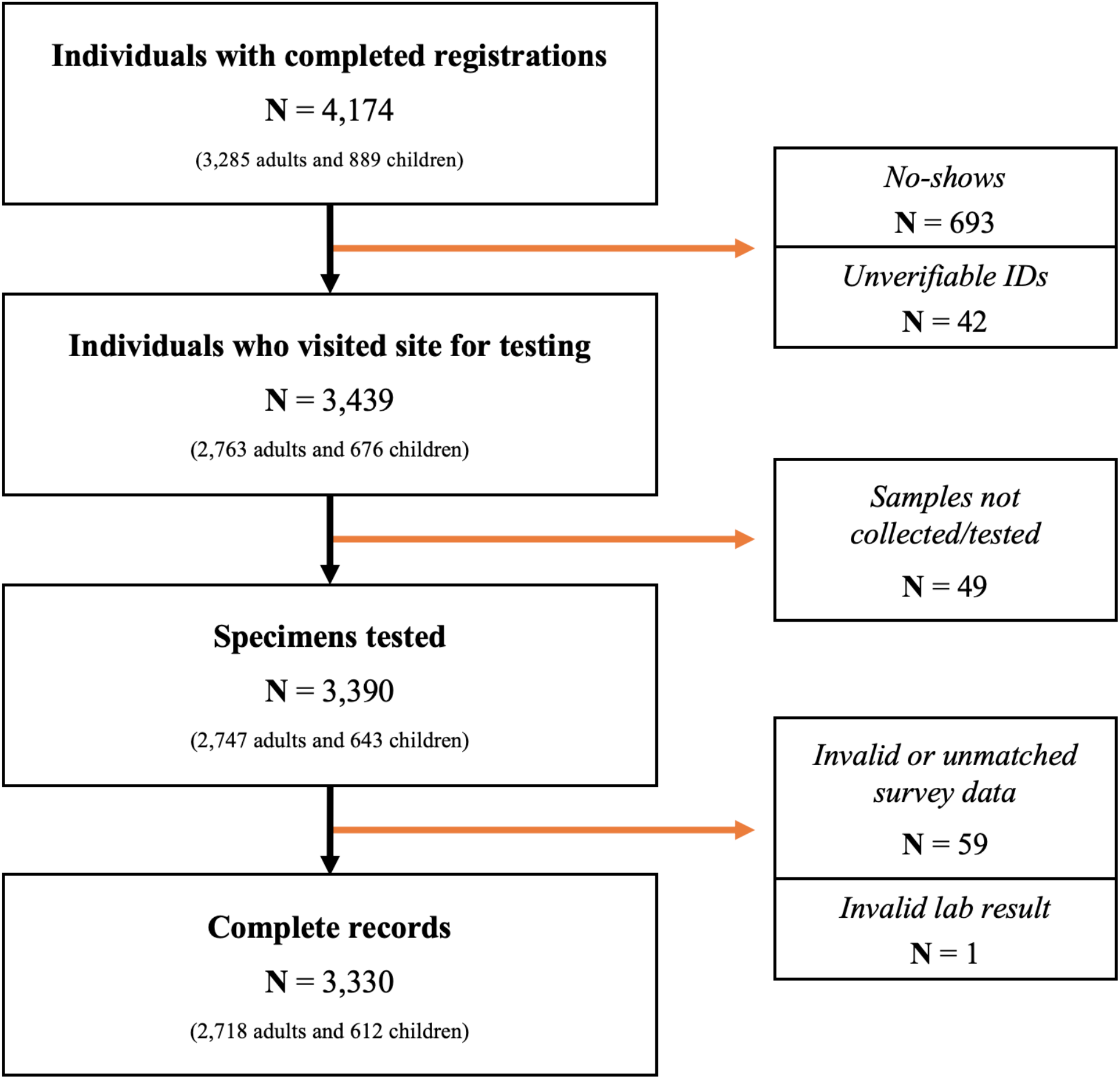
Of 3,439 individuals that arrived for testing, samples could not be obtained or tested (e.g. blood clotted) on 49, and an additional 60 were excluded because of invalid data (e.g. residence outside Santa Clara County), test data that could not be matched to a participant, or invalid test results. The final sample contained 3,330 records.

**Figure 2:**
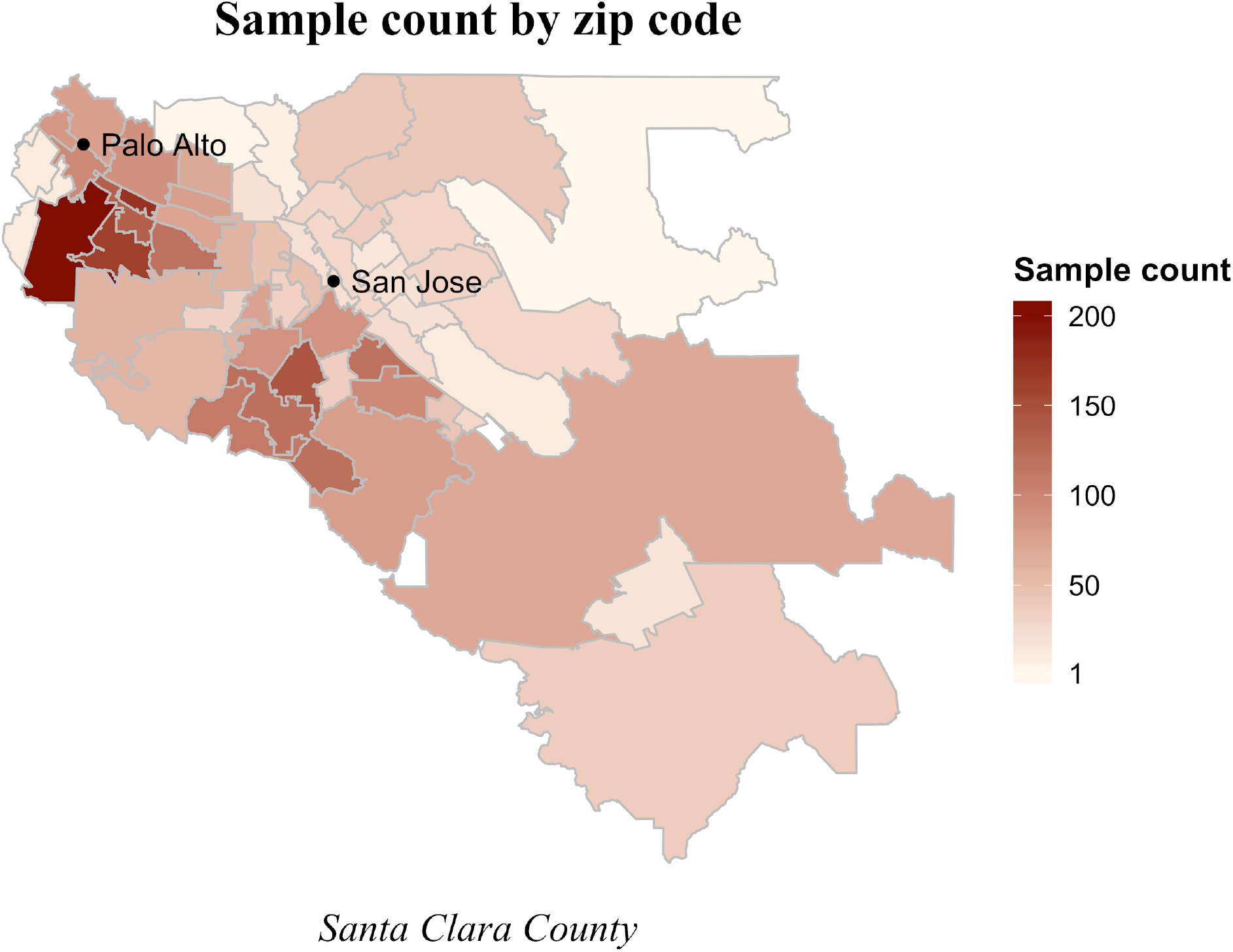
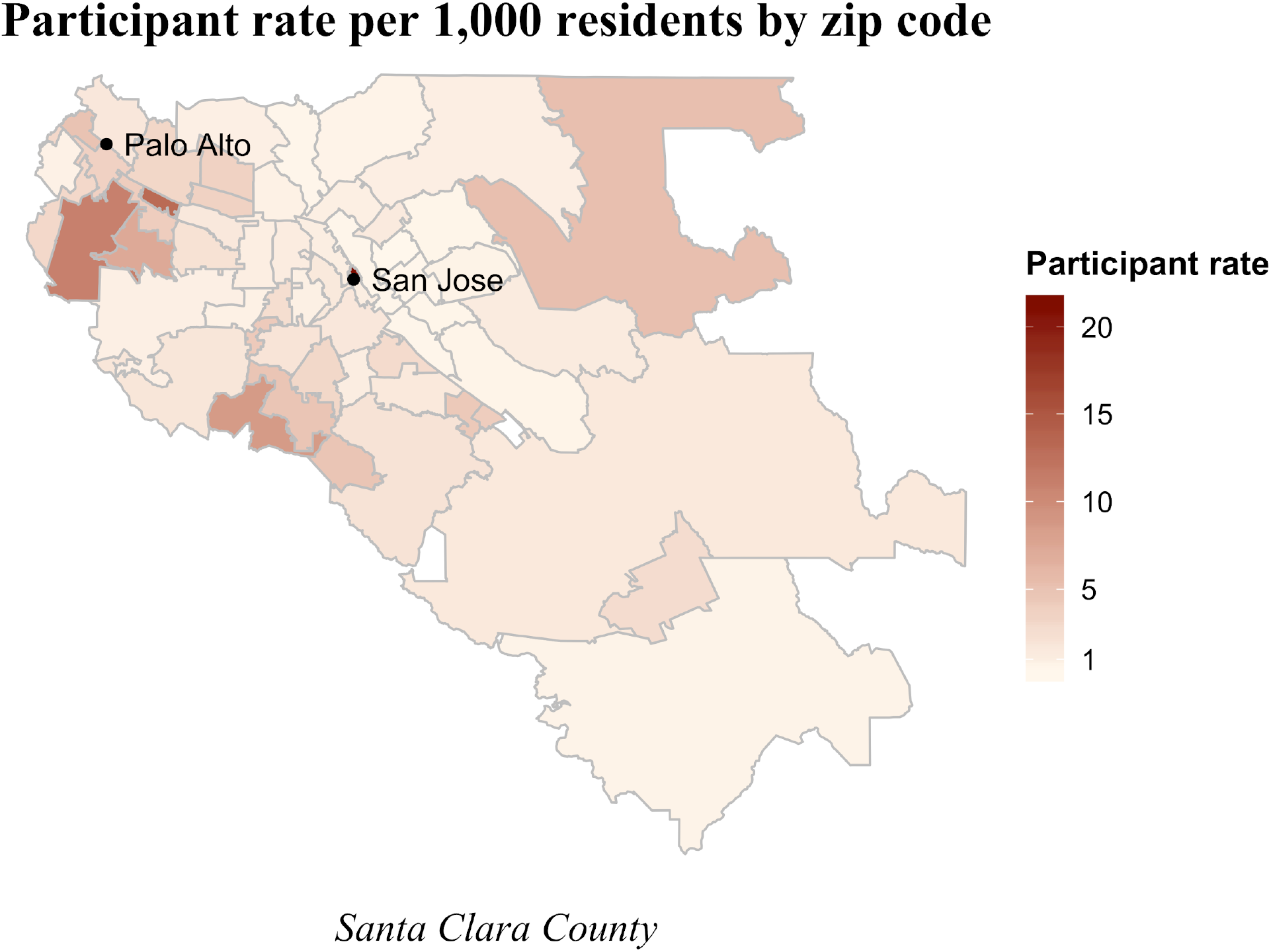
The number of registrations with complete records in our analytic dataset by zip code (panel A), and the participant rate per 1,000 residents in the zip code (panel B).

The total number of positive cases by either IgG or IgM in our unadjusted sample was 50, a crude prevalence rate of 1.5% (exact binomial 95CI 1.1-2.0%). Accounting for test sensitivity and specificity and sampling error, our point estimate of the unweighted population prevalence was 1.2% with a bootstrap 95% confidence interval of 0.7 to 1.8%. After weighting our sample to approximate Santa Clara County by zip, race, and sex, the prevalence was 2.8% (95CI 1.3 - 4.7%). Our confidence intervals on the weighted and unweighted sample prevalence account for three independent sources of uncertainty: sample prevalence, test sensitivity, and test specificity.

## Discussion

After adjusting for test performance characteristics and weighting for county demographics, we estimate that the seroprevalence of antibodies to SARS-CoV-2 in Santa Clara County in early April was 2.8%, with uncertainty bounds from 1.3% to 4.7% (unweighted prevalence 1.2%, 95CI 0.7-1.8%). These results represent the first large-scale community-based prevalence study in a major US county completed during a rapidly changing pandemic, and with newly available test kits. We consider our estimate to represent the best available current evidence, but recognize that new information, with more representative sampling and additional data on test kit performance, could result in updated estimates. Information indicating lower specificity would result in lower prevalence estimates. On the other hand, lower sensitivity, which has been raised as a concern with point-of-care test kits, would imply that the population prevalence would be even higher. New information on test kit performance should be incorporated as more testing is done, and we plan to revise our estimates accordingly.

The most important implication of these findings is that the number of infections is much greater than the reported number of cases. Using the weighted estimates, our data imply that, by April 1 (three days prior to the end of our survey) around 54,000 (95CI 25,000-91,000) people had been infected in Santa Clara County. The reported number of confirmed positive cases in the county on April 1 was 956, 55-fold lower than the number of infections predicted by this study (26-to-95-fold).^19^ This infection-to-case ratio, also referred to as an under-ascertainment rate, is meaningfully higher than current estimates.^10,20^ This ascertainment rate is a fundamental parameter of many projection and epidemiologic models, and is used as a calibration target for understanding epidemic stage and calculating fatality rates.^21,22^ The under-ascertainment for COVID-19 is likely a function of reliance on PCR for case identification which misses convalescent cases, early spread in the absence of systematic testing, and asymptomatic or lightly symptomatic infections that go undetected.

The extent of infection under-ascertainment is a central input for estimation of infection fatality rates from COVID-19. Many estimates of fatality rate use a ratio of deaths to lagged cases (because of duration from case confirmation to death), with an infection-to-case ratio in the 1-5-fold range as an estimate of under-ascertainment.^3,4,23^ Our study suggests that adjustments for under-ascertainment may need to be much higher.

We can use our prevalence estimates to approximate the infection fatality rate from COVID-19 in Santa Clara County. Through April 22, 2020, 94 people died from COVID-19 in the County. If our estimates of 54,000 infections represent the cumulative total on April 1, and we assume a 3 week lag from time of infection to death, up to April 22^24^, then 94 deaths out of 54,000 infections correspond to an infection fatality rate of 0.17% in Santa Clara County. If antibodies take longer than 3 days to appear, or if the average duration from case identification to death is less than 3 weeks, then the prevalence rate at the time of the survey was higher and the infection fatality rate would be lower. On the other hand, if deaths from COVID-19 are under reported or the health system is overwhelmed than the fatality rate estimates would increase. Our prevalence and fatality rate estimates can be used to update existing models, given the large upwards revision of under-ascertainment.

While our weighted prevalence estimate of 2.8% is indicative of the situation in Santa Clara County as of early April, other areas are likely to have different seroprevalence estimates based on effective contact rates in the community, variable effects of social distancing policies to date, and relative disease progression. The infection fatality rate in different locations may also vary and be substantially higher in places where the hospitals are overwhelmed (e.g. New York City or Bergamo), or where infections are concentrated among vulnerable individuals (e.g. populations without access to healthcare or nursing home residents). For example, in many European countries, 42-57% of deaths occurred in nursing homes and the same appears to be true for 25% of deaths in New York.^25^ Infection fatality rate estimates may be substantially higher in such settings.

Our prevalence estimate also suggests that, at this time, the large majority of the population in Santa Clara County remains without IgM or IgG antibodies to SARS-CoV-2. However, repeated serologic testing in different geographies, spaced a few weeks apart, is needed to evaluate the extent of infection spread over time.

This study has several limitations. The primary limitation concerns sample selection biases. Our sample may be enriched with COVID-19 participants, by selecting for individuals with a belief or curiosity concerning past infection. We discuss further and attempt to quantify the potential impact of this bias in the Additional Data and Response to Comments section. Our study may also have selected for groups of people more likely to skew our sample against COVID-19 participants. For example, our sample strategy selected for members of Santa Clara County with ready access to Facebook who viewed our advertisement early after the registration opened. Our sample ended up with an over-representation of white women between the ages of 19 and 64, and an under-representation of Hispanic and Asian populations, relative to our community. Those imbalances were partly addressed by weighting our sample by zip code, race, and sex to match the county. Our survey also selected for members of the population who were able to spare the time to drive to the testing site, which may have skewed our sample against essential workers. Our study was also limited in that it could not ascertain representativeness of SARS-CoV-2 antibodies in other communities with possibly high prevalence, such as homeless populations and nursing homes. The overall direction and magnitude of these selection effects are hard to fully bound, and our estimates reflect the prevalence in our sample, weighted to match county demographics.

The Premier Biotech serology test used in this study has not been approved by the FDA by the time of the study, and further validation studies for this assay are ongoing. We used existing test performance data to establish the test’s sensitivity and specificity. Additional validation of the assays used could improve further our estimates and those of other ongoing serosurveys. We note that the sole purpose of the kit in this study was to estimate prevalence, and not to provide information about individual antibody or immunity status.

Several teams worldwide have started testing population samples for SARS CoV-2 antibodies, with preliminary findings consistent with a large under-ascertainment of SARS CoV-2 infections. In early April, reports from the town of Robbio, Italy, where the entire population was tested, suggest at least 10% seropositivity;^26^ and data from Gangelt, a highly affected area in Germany,^27^ point to 14% seropositivity. Universal screening by PCR of women delivering in two New York hospitals (15% positive)^28^ and similarly screening of people in homeless shelters (36%-43% positive)^29,30^ also suggest potentially widely disseminated asymptomatic or minimally symptomatic infection in mid-April. On April 23, the first seroprevalence results were announced for New York, with 13.9% seroprevalence in the State and 21.2% in New York City, suggesting an under-ascertainment rate of 10 for New York.^31^ These data are compatible with our estimates since more testing per population was done in New York than in Santa Clara. A serosurvey in Los Angeles County, California on April 10 estimates seroprevalence of 4.1%.^32^ Our data from Santa Clara County suggest the spread of the infection is similar to other moderately-affected areas such as Los Angeles, but lower than areas with higher disease burden.

We conclude that based on seroprevalence sampling of a large regional population, the best estimates for the prevalence of SARS-CoV-2 antibodies in Santa Clara County were 2.8% by early April (95CI 1.3-4.7%). While this prevalence may be far smaller than the theoretical final size of the epidemic,^33^ it also suggests that the large majority of the population does not have antibodies and may be susceptible to the virus. Our prevalence and IFR estimates do not advocate for or refute the usefulness of any non-pharmaceutical interventions. Instead, these new data should allow for better modeling of this pandemic which can be used to better evaluate various interventions. It is important to note that the under-ascertainment rate in Santa Clara may change over time (e.g. depending on greater availability of testing) and the under-ascertainment rate may be different in other locations. Improved test accuracy and larger sample sizes with random sampling can further reduce uncertainty in estimates. Our work demonstrates the feasibility of seroprevalence surveys of population samples to inform our understanding of this pandemic’s progression, project estimates of community vulnerability, and monitor infection fatality rates in different populations over time. It may also be a useful tool for reducing uncertainty about the state of the epidemic, which carry public benefits.

## Data Availability

The data is not available for sharing at this time.

## Additional Data and Response to Comments

We received many questions and constructive comments in response to the 1^st^ version of our preprint. These included comments from colleagues within our institution with expertise in epidemiology and statistics as well as additional comments from hundreds of other scientists who offered critiques in public and/or in personal communications with our team. In this section we provide important updates and additional analyses that address the major issues with the first version of the preprint. This section includes new data and analyses; in addition, new data on test kit performance and the bootstrap estimation of the confidence intervals have been incorporated throughout. We welcome additional suggestions.

### Section 1: Test kit performance characteristics and implications for estimated population prevalence

In the 1^st^ version of the preprint manuscript we provide the following approaches to assessing the test kit performance:

a. Among 37 samples of known PCR-positive COVID-19 patients with positive IgG or IgM detected on a locally-developed ELISA test, 25 were kit-positive. Sensitivity 67.6% (95 CI 50.2-82.0% using exact binomial)
b. Among a sample of 30 pre-COVID samples, all 30 were negative. Specificity 100% (95 CI 90.5-100% using exact binomial with a simple transformation of the width of the Clopper-Pearson exact confidence interval to estimate uncertainty with p=1.0).
c. In the manufacturer’s data, among 75 samples of clinically confirmed COVID-19 patients with positive IgG, 75 were kit-positive, and among 85 samples with positive IgM, 78 were kit-positive. Sensitivity 91.8% (using the lower estimate based on IgM, 95 CI 83.8-96.6%).
d. In the manufacturer’s data, among 371 pre-COVID samples, 369 were negative. Specificity of 99.5% (95 CI 98.1-99.9%). (Since the initial publication, we learned that 368 were negative by either IgG or IgM, and we use the updated figure in the current manuscript.)
e. A combination of both data sources provides us with a combined sensitivity of 80.3% (95 CI 72.1-87.0%) and a specificity of 99.5% (95 CI 98.3-99.9%).

Since then, we have learned that, in (a), 27 of the specimens, not 25, were kit-positive, and that the specimens in (b) came from healthy adults collected at a hospital in New York. We have also received additional information about Premier Biotech test performance characteristics from multiple sources, including the manufacturer’s original data, test performance assessments for regulatory documents, and independent evaluations. The table below summarizes the information we received from each sample, including sample type and provenance.

| <u>Sample</u> | <u>Gold standard N</u> | <u>Kit agreement N</u> | <u>Sample type</u> | <u>Notes/provenance</u> |
| --- | --- | --- | --- | --- |
| ***Specificity (negative agreement)*** |  |  |  |  |
| 1 | 371 | 368 | Serum | Manufacturer's data based on pre-COVID-19 era sera |
| 2 | 30 | 30 | Serum | Pre-COVID-19 era healthy adults from New York |
| 3 | 70 | 70 | Serum | Adults tested negative for Lyme disease in a hospital in La Crosse Wisconsin pre-COVID-19 + adults admitted in early 2020 before first case in county |
| 4 | 1,102 | 1,102 | Serum | Adults admitted to hospitals pre-COVID-19 |
| 5 | 300 | 300 | Plasma | Pre-COVID-19 plasma donors, in heparin |
| 6 | 311 | 311 | Fingerstick blood | Healthy adults, company staff of Biotest, the kit manufacturer |
| 7 | 500 | 500 | Venous whole blood | Children and adults admitted to hospital pre-COVID-19, on heparin |
| 8 | 200 | 198 | Serum | Pregnant women pre-COVID-19 |
| 9 | 99 | 99 | Serum | Pre-COVID-19 serum of patients with <450 U/ml rheumatoid factor |
| 10 | 31 | 29 | Serum | Pre-COVID serum of patients with >600 U/ml rheumatoid factor |
| 11 | 150 | 146 | Serum | China CDC data collected from COVID-19-era hospital patients negative by PCR in Jiangsu province |
| 12 | 108 | 105 | Plasma | Pre-COVID-19 plasma donors |
| 13 | 52 | 50 | Serum | COVID-19-era PCR-negative specimens, some of which were positive for another respiratory virus |
| **Total** | **3,324** | **3,308** |  |  |
| ***Sensitivity (positive agreement)*** |  |  |  |  |
| 14 | 85 | 78 | Serum | Manufacturer's data using confirmed COVID-19 patients |
| 15 | 37 | 27 | Serum | Specimens with confirmed COVID-19 by PCR and IgG or IgM confirmed by ELISA |
| 16 | 35 | 25 | Serum | Specimens with confirmed COVID-19 by PCR (6-10 days from symptom onset) |
| **Total** | **157** | **130** |  |  |

Except for the samples 1, 12, 13, 14, and 16, we obtained individual results for all specimens in each sample.

We continue gathering information on test kit performance, and will incorporate as new data becomes available.

We use the pooled test performance based on the available information:

Sensitivity: 82.8% (exact binomial 95CI 76.0-88.4%)

Specificity: 99.5% (exact binomial 95CI 99.2-99.7%)

Of note, 3 of the negative control samples used for specificity calculations are from the COVID-19 era and thus have a chance that they may include some undiagnosed infections among these negative controls. Excluding these 3 datasets (datasets 6,11,13), the specificity is slightly higher (2801/2811, 99.6%).

There is some preliminary evidence that young patients with mild symptoms may have lower or even undetectable titers of antibodies than older patients.^34^ The sensitivity of the test kit was assessed based on samples from symptomatic patients who came to attention to be tested for SARS-CoV-2. If the sensitivity is lower in asymptomatic patients, then the prevalence may be under-estimated.

Finally, it needs to be stated that our kit only tests for the presence of IgG and IgM antibodies. The immune response to respiratory viruses is very complex and it involves multiple mechanism besides IgM and IgG antibodies, including IgA responses and other cellular mechanisms. For example, the antibody response to influenza in the upper respiratory track is dominated by IgA, and seroconversion in adults in terms of mucosal IgA responses seems to be higher than serum antibody-based seroconversion.^35^ IgA responses seem to be important also for SARS-CoV-2, and these were not captured by our kit.^36^ Further, while our current understanding of the test kit performance does not rule out the possibility of potential cross-reactivity with non-SARS-CoV-2 coronavirus strains, such as coronavirus HKU1, NL63, OC43, or 229E, the test kit’s high specificity across 3,324 SARS-CoV-2 negative samples suggests very low cross-reactivity given the relatively higher background prevalence of human coronavirus strains.^37^

### Section 2: Propagation of uncertainty about the specificity in estimation population prevalence

Several people had expressed concerns that, as the 95% CI on the specificity at the time of the release of version 1 was bounded at 98.3% on the low end, that implied that all our positives could in fact be false positives if there were a false positive rate of 1.7%. This is an intuitive stance, but deserves closer scrutiny. First, suggestions that the prevalence estimates may plausibly include 0% are hard to reconcile with documented data from Santa Clara (we had approximately 1,000 confirmed cases by PCR at the time of the survey) and what we know about the virus and its high contagiousness, as well as growing amounts of data from many other locations (such as Los Angeles and New York, as mentioned in the Discussion).

Second, additional data about the performance characteristics of the test kits is already helping improve the precision about the specificity point estimates, as noted above. With the additional kit performance data, the lower bound on the specificity 95CI is currently higher than in the original manuscript.

Third, for 0 true positives to be a possibility, one needs not only the sample prevalence to be less than (1-specificity) (something that is incompatible with the current 95CI or even 99CI of specificity), but also to have no false negatives (100% sensitivity) which is also entirely incompatible with the available data.

We maintain, as in the original manuscript, that “New information on test kit performance and population should be incorporated as more testing is done, and we plan to revise our estimates accordingly.”

Fourth, our Methods and Statistical Appendix go through our approach to carrying forward the uncertainty in the specificity, sensitivity, and the sample in estimating the uncertainty about the population prevalence. In particular, several commenters have argued that under a prior of low prevalence, the expected number of false positives in our sample exceeds the observed positives. Such an argument is misleading because our adjusted prevalence estimate is a combination of all 3 random variables. Our methods account for all three random variables simultaneously.

Finally, we now include the Statistical Appendix in the same file as the full paper for ease of reference.

To address the many constructive criticisms we have received regarding our standard error estimation – and in particular to address the point that our prior approach (the delta method) produces a symmetric confidence interval – we now calculate confidence intervals about our weighted and unweighted sample prevalence estimates based on a non-parametric percentile bootstrap that does not necessarily yield symmetric confidence intervals. Our bootstrap procedure resamples data from our actual datasets for sensitivity, specificity, and prevalence. We use the following procedure, repeated for each test performance scenario:

a. First, we draw a single bootstrap sample (i.e. with replacement) from the empirical distribution of the sensitivity data. For this sample, we calculate the mean value of sensitivity. Let 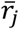 this value for the *j*^th^ iteration of this procedure. represent
b. We draw a single bootstrap sample from the empirical distributions of the specificity data. From this sample, we calculate the mean value of specificity. Let 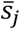 iteration of this procedure. represent this value for the *j*^th^
c. We draw a single clustered bootstrap sample from the Santa Clara County survey sample. That is, we draw clusters of individuals belonging to the same family with replacement. Each cluster either consists of a single adult, or an adult and child belonging to the same family. For our unweighted estimates, we calculate the unweighted fraction of individuals with either a positive IgG or IgM test reading. 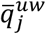 Let represent this value for the *j*^th^ iteration of this procedure.

For our weighted estimates, we run the same algorithm as above, except we replace step c with the following:

c’ We draw a single clustered bootstrap sample from the Santa Clara County survey sample. That is, we draw clusters of individuals belonging to the same family with replacement. Each cluster either consists of a single adult, or an adult and child belonging to the same family. For our weighted estimates, we calculate the weighted fraction of individuals with either a positive IgG or IgM test reading, calculated using the sample weights for each person in the bootstrap sample. Let 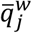 represent this value for the *j*^th^ iteration of this procedure. We recalculate the weights in each bootstrap sample using the same procedure we use for calculating our original sample weights: we recalculate sample weights in each bootstrap draw to align bootstrap zip/race/sex frequency with the county zip/race/sex population frequencies.
d We then calculate the test-error adjusted prevalence of antibodies to SARS-CoV-2 for the *j*^th^ bootstrap sample using the following formula (derived in the statistical appendix):

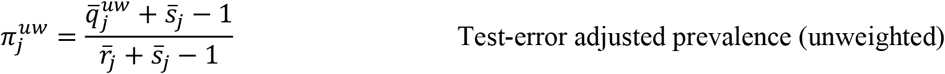

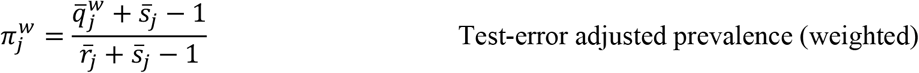
e Repeat the steps above for 10,000 bootstrap samples.
f In our tables, we report the 2.5^th^ percentile and the 97.5^th^ percentiles of the distributions of 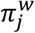 and 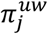 over the 10,000 bootstrap samples as the lower and upper ends of the 95% bootstrap confidence intervals. The point estimate we report are the respective means of these distributions. Of particular interest is the fact that even the minima of these distribution are above zero.

Our bootstrap analysis shows the following estimates (also in Table 2):

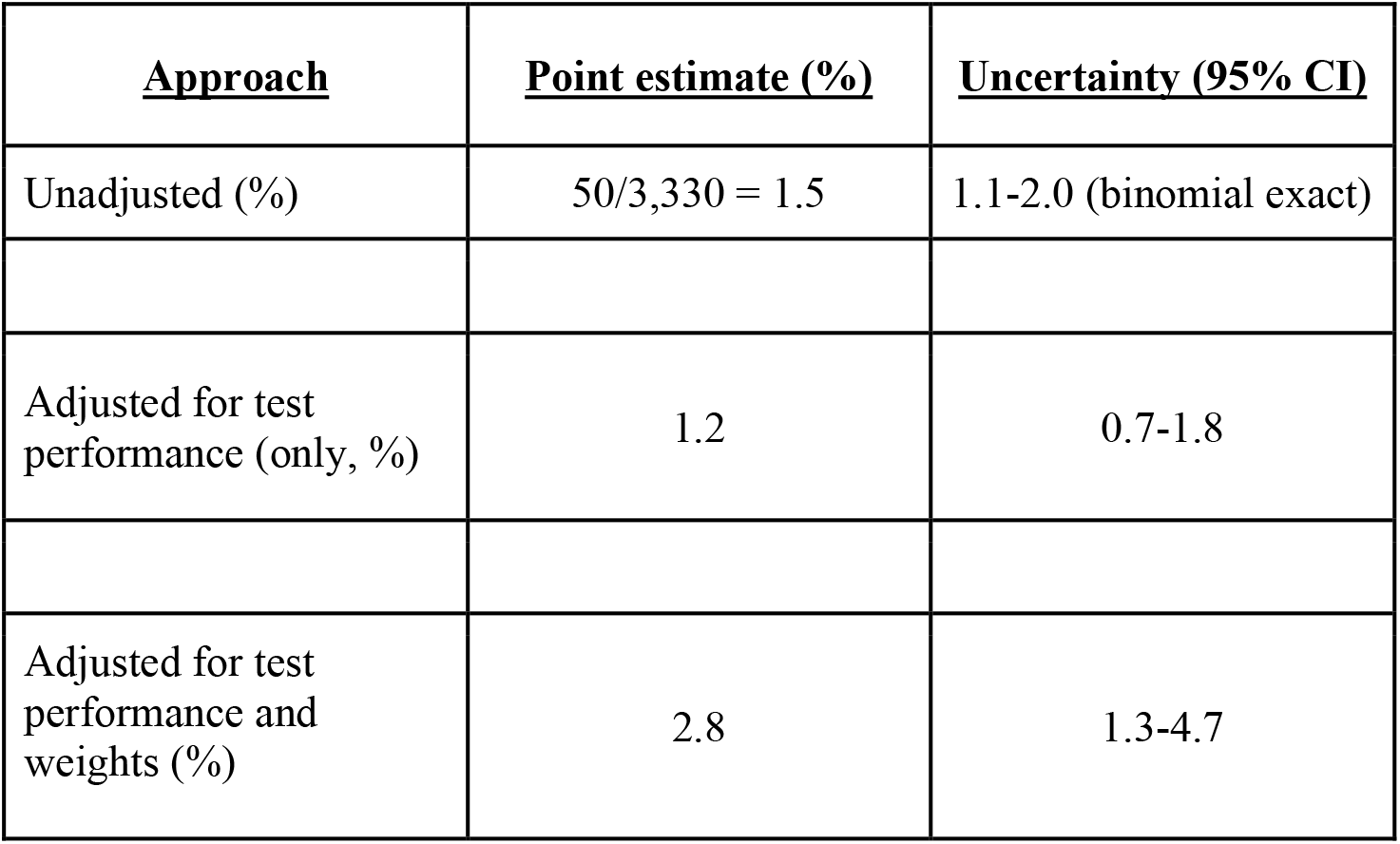

**Table 2:**
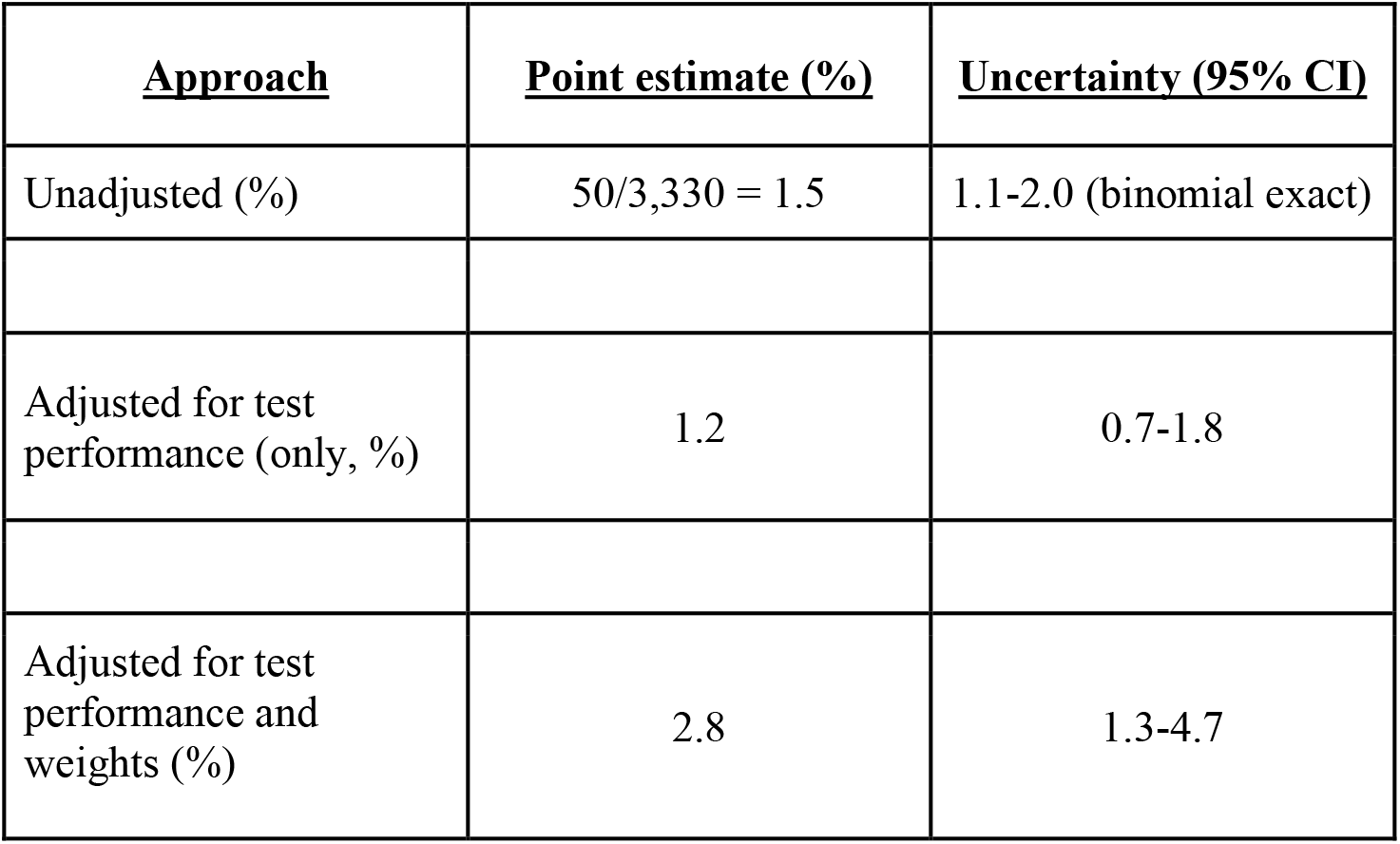
Prevalence estimation in Santa Clara County. We report the prevalence and uncertainty bounds of estimates from unadjusted frequency counts, estimates adjusted for test performance characteristics, and estimates adjusted for test performance characteristics and weighted by zip code, race/ethnicity, and sex. We estimate uncertainty using the bootstrap as described in Methods and below.

**Table 3:**
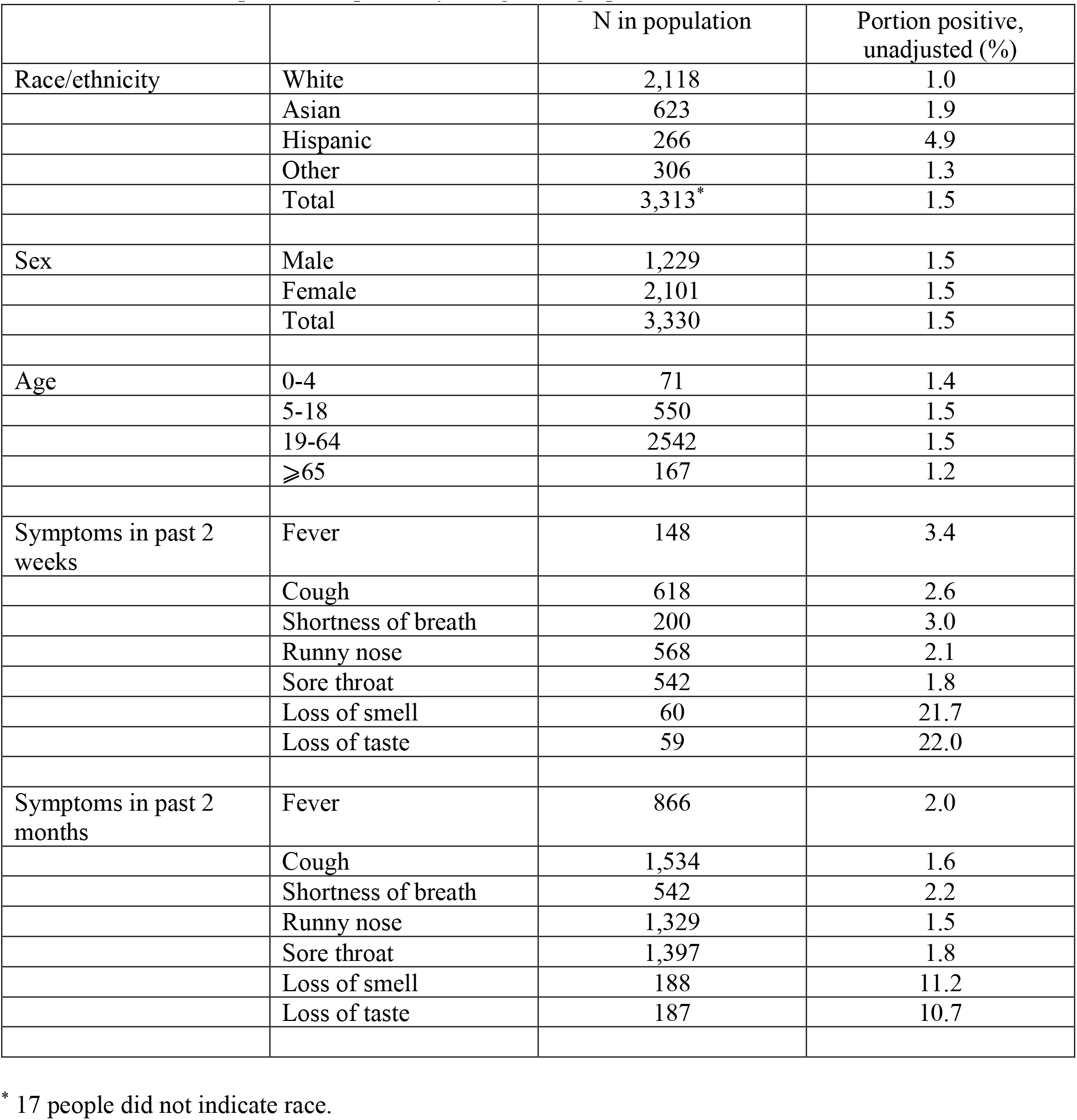
Univariate frequencies of positivity along demographic and clinical features

### Section 3: Sample bias in favor on those with higher likelihood of COVID-19

Several commentators who read the preprint expressed a concern that our sample was enriched in people who had COVID-19 because those who thought they may have had COVID-19 would have greater desire to get tested and register for the study. While this is a limitation of any study involving voluntary participation, this is a source of bias that we assessed in several ways.

First, we provide additional narrative detail about the participant recruitment process. The Facebook ad was designed to reach individuals determined by Facebook’s algorithm to reside in Santa Clara County zip codes, with representative reach across zip codes. Within 2 hours from the moment our Facebook ad went live on April 2, we had nearly 1,000 registrations, dominated by relatively wealthy zip codes. These registrations appear to have been driven by sharing of the Facebook ad and registration links in local social media and listservs amongst residents of relatively wealthy zip codes. We learned of at least one incident where the survey link from the Facebook ad, as well as inaccurate information about the study purpose, were shared to a listserv without our knowledge or consent. During the study, when we realized that our registrations were heavily skewed towards those zip codes, we moved to preferentially accept registrations from under-represented zip codes of the county into the sample. While we acknowledge the limitations of this approach to participant recruitment, we were also balancing a quick timescale (attempting to recruit >2,500 participants within 24 hours) and current shelter-in-place policies. Alternative convenience sampling approaches, such as sampling outside of grocery stores (an approach other seroprevalence studies are using), would have captured individuals from a smaller subset of zip codes and been subject to other selection concerns.

As has been suggested, it is possible that across all populations there was a bias towards those with greater suspicion for current or past disease. We believe the speed with which registrations happened more likely selected for those who were ready to respond to a notice on Facebook. At the end of the ∼24 hours when we had the registration open, we had ∼11,000 people get to the registration site from Facebook alone, suggesting that the most eager participants who managed to sign up quickly entered our sample, not necessarily those at greatest risk. During the survey days, we were concerned about the apparent over-representation of wealthy and healthy-appearing participants. This bias is consistent with a common type of healthy volunteer bias which leads to under-participation of sicker populations in trials and surveys.

Second, we performed the following analyses to assess the possible magnitude of selection bias. We asked our participants about cough and fever symptoms in the past 2 weeks and in the past 2 months. We use this information as follows. First, we assess the frequency of symptoms in our sample:

|  | Unweighted sample (n, %) |
| --- | --- |
| Cough + fever in past 2 weeks | 101, 3.0 |
| Cough + fever in past 2 months | 692, 20.8 |

We then assess the relationship of symptoms to test positivity using simple frequencies. The table below shows that symptoms are positively associated with testing positive. (This is, incidentally, indirect evidence that the people who tested positive in our sample are true positives rather than false positives.)

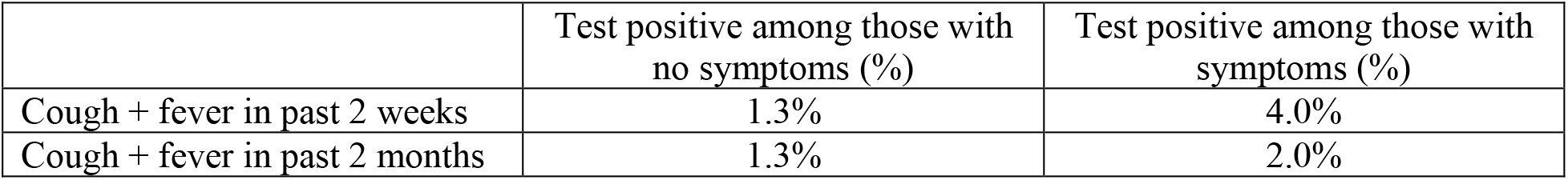

We then use these estimates to assess the impact of potential sample enrichment relative to the background prevalence of COVID-19-related symptoms in the county. We do this by estimating the number of additional positive cases in our sample relative to a scenario where our sample would have three times fewer people with cough and fever (chosen to approximate a lower bound). In other words, we calculate the population prevalence under two scenarios: (S1) where 1.0% of our sample came in with cough and fever in past two weeks (a third of 3.0%) or (S2) where 6.9% of our sample came in with cough and fever in past two months (a third of 20.8%). We calculate the prevalence as in our main manuscript, using the bootstrap, adjusted for test performance characteristics, for each scenario. For example, in scenario S1, the 2.0% decline in cough and fever symptoms (3.0% to 1.0%) would imply about 67 fewer people with cough and fever symptoms (3,330 × 2.0%). We then multiply this with the marginal probability of cough and fever symptoms on testing positive (2.7 percentage points), which implies 2 fewer positives. Similarly, scenario S2 implies 4 fewer positives. The implications of this potential bias for the weighted and test-performance adjusted prevalence is below:

|  | Additional positives implied by potential sample bias | Population prevalence in case-depleted sample (%; 95CI) |
| --- | --- | --- |
| Cough + fever in past 2 weeks | 2 | 2.7, (1.3-4.6) |
| Cough + fever in past 2 months | 4 | 2.6, (1.1-4.5) |

The adjusted and weighted prevalence in our original sample was 2.8%. In other words, under assumptions of 3-fold selection based on fever and cough, our population prevalence estimates are only moderately different from our base estimates. Moreover, for February and March, the proportion of participants with cough + fever may actually be lower than what is typical in California, and, if so, the bias in the estimation of prevalence may be even in the opposite direction (prevalence may be a bit underestimated).^38^

Weighting our sample to account for differences relative to the demographics of Santa Clara County plays an important role in this study. Applying weights to our sample results in prevalence estimates that are higher than the unweighted sample. Below we provide the distribution of the weights we applied to our sample, separately for those that tested positive (orange) and negative (green). The figure again illustrates that those that tested positive were, on average, drawn from demographic groups that were under-represented in our sample, while those that tested negative were, on average, drawn from groups that were over-represented in the sample.

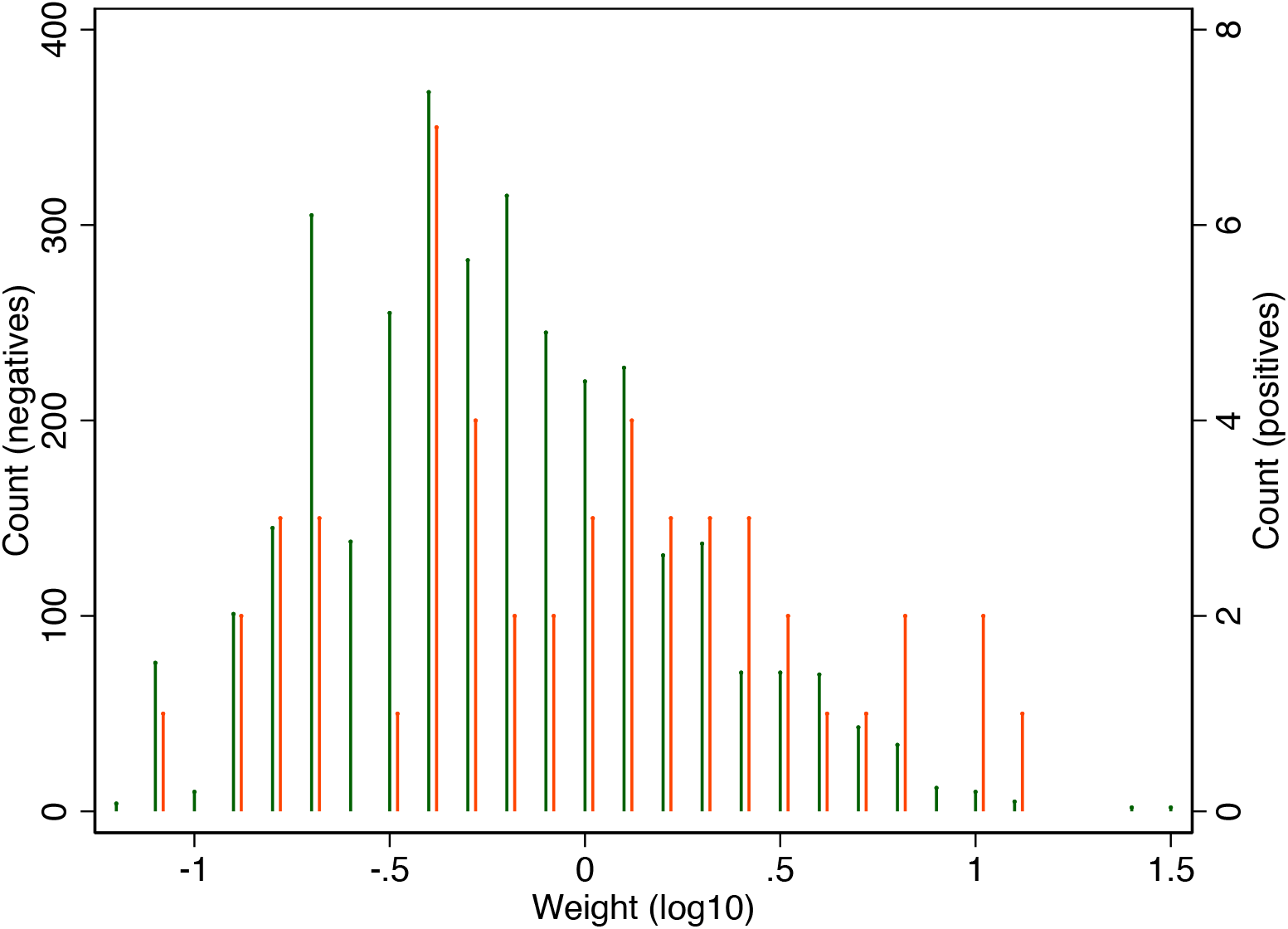

### Section 4: Univariate frequencies of positivity based on demographics and symptoms

Multiple readers asked about the relative frequency of antibody positivity along demographic features and symptoms. This was not provided in the original manuscript to allow a focus on the population prevalence. However, we provide this table here. While any gradients may be spurious, the differences in positivity along race/ethnicity and symptoms that have been observed to go along with COVID-19 may be taken as supportive of the ability of our test kit to distinguish true positives and true negatives.

We are grateful for the enormous interest in our work and for the helpful suggestions raised by other scientists that have helped us strengthen our data and inferences. We feel that our experience offers a great example on how preprints can be an excellent way of providing massive crowdsourcing of helpful comments and constructive feedback by the wider scientific community in real time for timely and important issues. We continue to strive for transparency in our approaches to dealing with these issues and with interpreting the results. We are open to all debates over this issue. Ultimately, this is just one study, and more studies will further our understanding of the seroprevalence of SARS-CoV-2 antibodies in Santa Clara and other counties and locations around the world.

## Statistical Appendix

In this appendix, we describe our statistical methods in more detail. Section 1 describes our approach to calculating population weights. Section 2 describes our approach to adjusting our population prevalence estimate for the sensitivity and specificity properties of the test kit we are using.

### Population weighting

In all but the unadjusted prevalence results, we reweight our sample to reflect the sex, race/ethnicity, and zip code of residence distribution of Santa Clara County. We derive these weights from the 2018 American Community Survey, from which we derived an estimate of the population of each zip code, as well as the race/ethnicity and sex distribution of county residents. We applied the county-wide sex distribution to each race/ethnicity group in each zip code to estimate the number of people within each zip-race-sex group. For example, zip code 95037 has a total of 51,652 residents, of which 50.6% are female, 49.7% are white, 7.7% are Asian, 33.0% are Hispanic, and 9.5% are other. We applied the female proportion to each race category in the zip code to obtain the number of residents in each zip-race-sex group.

Let *Epop*_*zip,race,aex*_ be the number of people in each zip-race-sex cell produced by this calculation. Let *SmpSz*_*zip,race,aex*_ be the number of people in our sample population in each cell. We need to up-weight cells that are underrepresented in our population relative to their frequency in Santa Clara County, and down-weight cells that are overrepresented. We can accomplish this by weighting proportional to the following ratio:

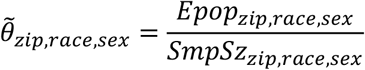

We renormalize the weights so that the sum of our weights equals the size of the SCC sample, *N*. Define

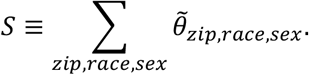

Our final sample weights are:

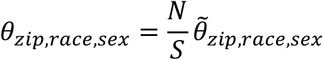

This formula provides sample weights that are the same for all individuals in our sample within each zip/race/sex group. To make our notation clear, let *i* denote each individual in our population and let *zip*_*i*_, *race*_*i*_, and *sex*_*i*_ be the zip code of residence, race, and sex of individual *i*. We assign a sample weight of 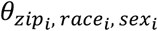 for person *i*.

To provide a concrete example, suppose the county had two zip codes, A and B. The populations of each zip code include 10,000 men and 10,000 women (40,000 total population). Our sample included 250 men and 500 women from zip A, and 750 men and 1500 women from zip B. This is typical of the imbalance in our sample. Applying the formula above, we get relative weights of 3 each man in zip A, 1.5 for each woman in zip A, 1 for each man in zip B, and 0.5 for each woman in zip B.

### Adjusting the prevalence estimate for test kit accuracy

Our main goal is to derive an estimate of the prevalence of specific COVID-19 antibody seroconversion in Santa Clara County. However, we observe an inaccurate measure of antibody presence because the test kit we use has both false positives and false negatives.^39^ In this section, we describe our approach to adjusting for these errors. To spare notation, we do not incorporate the sample weighting process we describe above. Introducing sample weighting would complicate our notation, but would not change the approach. The analytic weights were used in the results shown in Table 2.

Let *Π* = *P*(*COVID* +) represent the population prevalence of antibodies to COVID-19, and let *q* = *P*(*TEST* +) be the proportion of participants who test positive in our sample (this latter quantity measured using our sample weights). Note: we consider *TEST* + as any band on the test kit indicating the presence of IgG or IgM antibodies or both.

Let *r* = *P*(*TEST* + | *COVID* +) be the sensitivity of the test and let *s* = *P*(*TEST* − | *COVID* −) be the specificity of the test. Let *z* = *P*(*COVID* + | *TEST* +) be the positive predictive value of the test, and *y* = *P*(*COVID* + | *TEST* −) be the (one minus) the negative predictive value of the test.

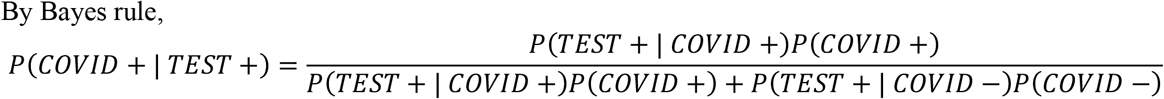

and

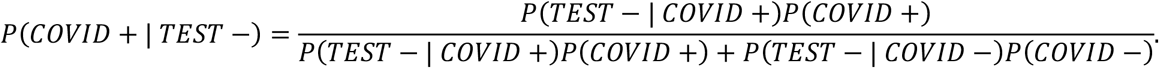

Rewriting these in our notation, we have:

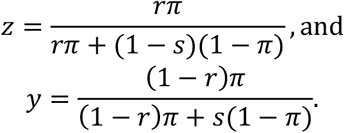

By the definition of conditional probability, we also have:

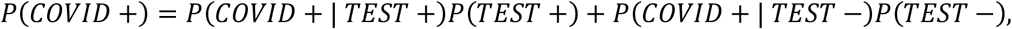

Or in our notation:

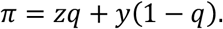

If we plug in our expressions for *y* and *z* and simplify, we have a quadratic expression in *Π*.

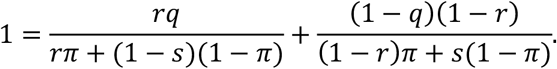

We solve for *π* as a function of the sample prevalence, sensitivity, and specificity:

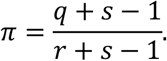

There is one important caveat to this formula: it only holds as long as (one minus) the specificity of the test is higher than the sample prevalence. If it is lower, all the observed positives in the sample could be due to false-positive test results, and we cannot exclude zero prevalence as a possibility. As long as the specificity is high relative to the sample prevalence, this expression allows us to recover population prevalence from sample prevalence, despite using an imperfect diagnostic test.

## Article Information

### Corresponding author

Eran Bendavid

